# Mechanical Versus Manual Ventilation During Cardiopulmonary Resuscitation: A Systematic Review and Meta-Analysis

**DOI:** 10.64898/2025.12.19.25342720

**Authors:** Gunaseelan Rajendran, Sasikumar Mahalingam, Anitha Ramkumar, Gayathri Pandy, Ezhilkugan Ganessane, Vijayanthi Vijayan, Purushothaman Ranagasamy, Hanumantha Rao

## Abstract

**Background:** Manual bag-valve ventilation during cardiopulmonary resuscitation (CPR) is prone to substantial variability in tidal volume and respiratory rate, frequently resulting in hyperventilation. The clinical effectiveness of mechanical ventilation as an alternative strategy remains uncertain.

**Objectives:** This systematic review and meta-analysis compared mechanical versus manual ventilation during adult CPR to assess return of spontaneous circulation (ROSC), survival to hospital discharge, and neurological outcomes.

**Methods:** We searched PubMed, Embase, and Scopus (inception through October 2025) for randomized controlled trials and observational studies comparing mechanical and manual ventilation during adult CPR. We conducted separate meta-analyses for randomized trials and observational studies using random-effects models and assessed evidence certainty using GRADE methodology. Primary outcomes were ROSC, survival to discharge, and favorable neurological outcome (Cerebral Performance Category 1–2).

**Results:** Eight studies (5,130 patients) met inclusion criteria. Mechanical ventilation was associated with higher ROSC (odds ratio [OR] 1.22; 95% confidence interval [CI] 1.07–1.38; p=0.002; I²=8%), survival to discharge (OR 1.39; 95% CI 1.08–1.77; p=0.009; I²=0%), and favorable neurological outcome (OR 1.61; 95% CI 1.04–2.48; p=0.03; I²=0%) compared with manual ventilation. In randomized trials (n=120), mechanical ventilation showed a trend toward improved ROSC (OR 1.49; 95% CI 0.73–3.07; p=0.27) but lacked statistical significance. Observational studies (n=7,081) demonstrated an association between mechanical ventilation and higher ROSC (OR 1.21; 95% CI 1.03–1.42; p=0.02; I²=34%). Post-ROSC arterial blood gases showed improved oxygenation (mean difference 13.01 mmHg higher pO₂ ; p<0.0001) and lower pCO₂ levels (mean difference 15.12 mmHg lower; p<0.00001) with mechanical ventilation. GRADE assessment indicated low-certainty evidence for clinical outcomes and moderate-certainty evidence for physiological outcomes.

**Conclusions:** Mechanical ventilation during CPR was associated with higher rates of ROSC, survival, and favourable neurological outcomes, along with more controlled post-ROSC physiological parameters. However, the certainty of evidence is low, driven largely by confounded observational data and limited randomized trial evidence. These findings are hypothesis-generating and should not be interpreted as causal. Confirmation in adequately powered randomized controlled trials is required before changes to practice or guidelines can be recommended.

**WHAT IS NEW?:**

- This systematic review and meta-analysis, stratified by study design, synthesizes the available evidence comparing mechanical and manual ventilation during adult cardiopulmonary resuscitation across eight studies involving 5,130 patients.
- Mechanical ventilation was associated with higher rates of return of spontaneous circulation, survival to hospital discharge, and favorable neurological outcome compared with manual ventilation; however, these associations are derived largely from observational data with low certainty of evidence.
- Mechanical ventilation demonstrated more consistent post-resuscitation arterial blood gas parameters—higher oxygenation and lower carbon dioxide levels—suggesting physiologic benefits, although these findings also require confirmation in randomized trials.

**CLINICAL IMPLICATIONS?:**

- Mechanical ventilation may offer a more standardized approach to delivering tidal volumes and respiratory rates during CPR, potentially mitigating the variability and risk of hyperventilation inherent to manual bag-valve ventilation.
- Because the evidence supporting improved clinical outcomes is low certainty and primarily observational, the observed associations should not be interpreted as causal. These results are hypothesis-generating and highlight an important area for further investigation rather than indicating definitive clinical benefit.
- If mechanical ventilation is used during CPR, implementation should prioritize protocolized ventilator settings (e.g., tidal volume 6–7 mL/kg and respiratory rate 10 breaths/min) and strict adherence to high-quality chest compressions.
- Adequately powered randomized controlled trials are needed to determine whether mechanical ventilation confers true clinical benefit and to inform future guideline recommendations.

## **1.** Introduction

Manual bag-valve ventilation remains the predominant method of airway management during cardiopulmonary resuscitation (CPR),^1^ yet its delivery is highly inconsistent in real-world practice. Providers frequently administer excessive ventilation rates, variable tidal volumes, and poorly coordinated breaths, leading to hyperventilation during resuscitation.^2–5^ These deviations increase intrathoracic pressure, impede venous return, and diminish coronary and cerebral perfusion pressures, which are physiologic consequences known to reduce the probability of return of spontaneous circulation.^6–9^ Variability is further amplified by provider fatigue, differences in clinical experience, and the demands of multitasking during high-acuity CPR. Although contemporary resuscitation guidelines caution against hyperventilation, they offer limited practical direction on achieving consistent ventilation and provide no specific recommendations regarding the use of mechanical ventilators during active CPR.^1^ As a result, mechanical ventilation is increasingly adopted to standardize ventilation and reduce provider workload, yet its clinical effectiveness remains uncertain.

Both manual bag-valve (BV) ventilation and mechanical ventilation carry inherent advantages and limitations. BV devices are lightweight, portable, and simple to operate, but they typically lack real-time feedback on delivered pressures, volumes, or rates, making them prone to inconsistent or potentially harmful ventilation—especially given providers’ tendency to hyperventilate during CPR, which can reduce coronary perfusion and cardiac output.^10–12^ Mechanical ventilators mitigate this variability and can free an additional provider for other resuscitation tasks, yet they introduce challenges such as breath–compression asynchrony, fluctuating chest wall compliance, and resultant high-pressure cutoffs that may yield suboptimal tidal volumes.^13,14^ These differences in breath delivery and minute ventilation between manual and mechanical techniques may ultimately influence oxygenation during resuscitation. Studies comparing mechanical and manual ventilation during CPR have been limited in number and have yielded inconsistent findings. The available evidence is heterogeneous in design and quality. Importantly, this fragmented body of evidence has not been systematically synthesized. The overall impact of mechanical ventilation during CPR remains uncertain.

Given the growing clinical use of mechanical ventilation during CPR and the absence of a comprehensive evidence synthesis, a formal evaluation of clinical outcomes is needed. This systematic review and meta-analysis aimed to compare mechanical ventilation versus manual bag-valve ventilation during adult CPR by analyzing return of spontaneous circulation, survival to hospital discharge, and favorable neurological outcome. To enhance clarity and interpretability, we conducted separate meta-analyses for randomized controlled trials (RCTs), nonrandomized studies (nRCTs), and the overall pooled dataset. The certainty of evidence for each analysis—RCTs, nRCTs, and the combined evidence—was evaluated independently using the GRADE framework.

## 2. METHODS

### 2.1 Protocol and Registration

This systematic review and meta-analysis was conducted in accordance with the *Preferred Reporting Items for Systematic Reviews and Meta-Analyses (PRISMA) 2020* and *Meta-analysis of Observational Studies in Epidemiology (MOOSE)* guidelines.^15,16^ The review protocol was prospectively registered in the International Prospective Register of Systematic Reviews (PROSPERO; registration ID CRD420251169035; registered October 15, 2025).

### 2.2 Search Strategy

A comprehensive electronic search was performed across three databases namely *PubMed/MEDLINE*, *Embase* and *Scopus* from inception to October 2025. The search combined controlled vocabulary and free-text terms related to “*cardiac arrest*,” “*cardiopulmonary resuscitation*,” “*mechanical ventilation*,” and “*manual ventilation”* using Boolean operators. No language restrictions were applied.

Additional studies were identified through backward and forward citation searches (snowballing method) of relevant articles and reference lists, as well as manual searches of major resuscitation conference abstracts. Authors of eligible studies were contacted for clarification or missing data where necessary. The complete search strings for each database are provided in Supplemental Table S1. All records were imported into Rayyan (Qatar Computing Research Institute) for duplicate removal and screening.^17^

### 2.3 Eligibility Criteria

Studies were included if they met the following criteria:

- **Population:** Adults (≥18 years) experiencing in-hospital or out-of-hospital cardiac arrest and undergoing active CPR.
- **Intervention:** Mechanical ventilation during CPR, including modes such as intermittent positive-pressure ventilation (IPPV), chest compression-synchronized ventilation (CCSV), or automatic transport ventilators.
- **Comparator:** Manual ventilation using a bag-valve-mask (BVM) or resuscitator bag.
- **Outcomes:**

- *Primary outcomes:* The outcomes of interest were: <u>Return of spontaneous circulation (ROSC):</u> Any return of a palpable pulse during or after CPR (as defined by the original studies). <u>Survival to hospital discharge:</u> The proportion of patients who were alive at discharge from the hospital. <u>Favorable neurological outcome:</u> Defined as a good functional outcome at discharge (Cerebral Performance Category [CPC] score of 1 or 2).^18^ Where studies reported multiple time points, we extracted outcomes at the time of hospital discharge or closest follow-up.
- *Secondary outcomes:* Arterial or venous blood-gas parameters during CPR and adverse events (barotrauma, hyperventilation syndromes).
- **Study design:** Randomized controlled trials (RCTs) and non-randomized studies.

Studies were excluded if they (1) involved pediatric patients (<18 years), (2) used simulation, animal, or manikin models, (3) addressed ventilation only after ROSC, or (4) were reviews, editorials, or case reports.

### 2.4 Study Selection and Data Extraction

Two reviewers (GR, SM) independently screened titles and abstracts of all retrieved records for potential eligibility. The full texts of all potentially eligible articles were then obtained and assessed in duplicate against the inclusion criteria. Discrepancies between reviewers were resolved by discussion and consensus, with a third reviewer (AR) adjudicating unresolved cases.

For each included study, two reviewers (GR, SM) independently extracted data using a standardized data collection form. Extracted information included study characteristics (authors, year, setting, design), patient characteristics (age, out-of-hospital vs in-hospital arrest), intervention details (type of mechanical ventilator, timing of ventilation), and comparator details (BVM technique). We also recorded the number of participants randomized or enrolled, and the number of events for each outcome. Data were collected for ROSC, survival to hospital discharge, and favorable neurological outcome as defined below. Any discrepancies in data extraction were resolved by discussion with a third reviewer (AR) and verification against the source. If outcome data were unclear or missing, study authors were contacted for clarification.

### 2.5 Risk of Bias Assessment

Risk of bias for randomized controlled trials was assessed using the Cochrane Risk-of-Bias 2 (RoB 2) tool for each outcome, and for non-randomized studies using ROBINS-I frameworks.^19,20^ Two reviewers (GR, AR) performed independent assessments, with disagreements resolved by consensus with third reviewer (SM). Studies were categorized as having low, some, or high risk of bias. The impact of study quality on pooled estimates was explored through sensitivity analyses excluding high-risk studies. Risk-of-bias assessments are summarized in Supplemental Figure S2. The risk-of-bias plot was generated using shinyapps, developed by McGuinness and Higgins.^21^

### 2.6 Statistical Analysis

Statistical analyses were conducted when at least two studies reported comparable outcome data. For dichotomous outcomes, pooled odds ratios (ORs) with 95% confidence intervals (CIs) were calculated. Because traditional DerSimonian–Laird (DL) random-effects models may underestimate between-study variance when the number of included studies is small, we used the restricted maximum likelihood (REML) method for all primary random-effects analyses. In addition to frequentist analyses, a Bayesian meta-analysis using Bayesian Model Averaging (BMA) was performed to provide a probabilistic assessment of treatment effects and to account for uncertainty between fixed- and random-effects models. We applied weakly informative priors, using a Normal (0,1) prior for the pooled log OR to reflect equipoise and a Half-Normal (0,0.5) prior for between-study heterogeneity (τ), consistent with recommendations for sparse meta-analytic datasets. Posterior distributions, inclusion probabilities, Bayes factors (BFs), and 95% credible and prediction intervals were estimated via Markov Chain Monte Carlo sampling. Bayes factors were interpreted according to established thresholds, with BF values between 1 and 3 considered “anecdotal” evidence for a non-zero effect, indicating only weak support over the null model. The Bayesian analysis was included to complement the REML results by enabling probability-based interpretation of effect estimates, stabilizing inference with priors in the context of limited data, and providing prediction intervals to characterize uncertainty in future studies. Continuous outcomes were synthesized using mean differences or standardized mean differences, as appropriate.

Random-effects models were chosen a priori for all analyses to account for anticipated clinical and methodological heterogeneity across studies, including differences in ventilation modes, arrest setting, and airway strategies. Fixed-effects models were not used for primary analyses. Meta-analyses were performed separately for randomized controlled trials and nonrandomized studies, followed by an overall pooled analysis that combined all eligible study designs. Statistical heterogeneity was assessed using Cochran’s Q test (α = 0.10) and quantified with the I² statistic.

Subgroup analyses were planned for the location of cardiac arrest, categorized as out-of-hospital versus in-hospital arrest. Publication bias was evaluated using Egger’s regression test when at least ten studies were available for a given outcome. Statistical significance was defined by a two-tailed P value less than 0.05.

All statistical analyses were performed using STATA version 17 (StataCorp, College Station, Texas) and Review Manager version 5.4 (Cochrane Collaboration).

## 3 Results

### 3.1 Study Selection

A total of 2,624 citations were identified through systematic database searches (PubMed: n = 1,308; Embase: n = 701; Scopus: n = 615). After removal of 413 duplicates using *Rayyan* software, 2,211 unique records were screened by title and abstract. Of these, 90 full-text articles were assessed for eligibility. Following full-text review, 8 studies met the inclusion criteria and were included in the final systematic review and meta-analysis (Figure 1).^22–29^

**Figure 1.**
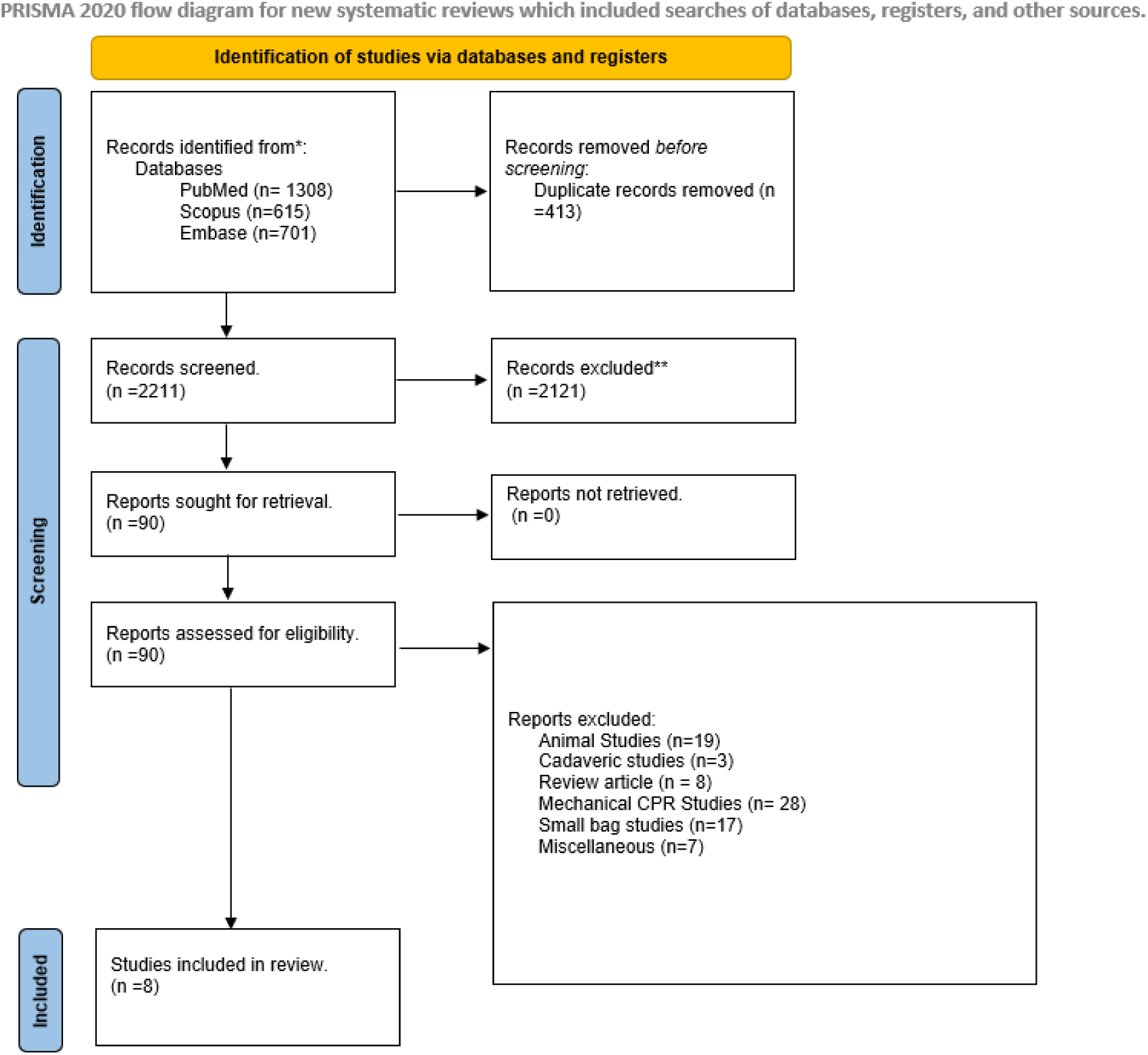
PRISMA Flow Diagram of Study Selection. Flow diagram summarizing the identification, screening, eligibility assessment, and final inclusion of studies in the systematic review and meta-analysis. A total of 2,624 records were identified, 213 duplicates removed, 2,211 titles/abstracts screened, 90 full texts reviewed, and 8 studies met inclusion criteria.

The most common reasons for study exclusion were animal or cadaveric models (n = 22), studies involving mechanical chest compression devices rather than ventilation (n = 28), small experimental “bag studies” during CPR studies (n = 17), narrative reviews (n = 8), and other miscellaneous non-comparative reports (n = 7). Importantly, outcome reporting was not uniform across studies: (a) Eight studies were included in the review overall. (b) Seven studies provided extractable data for return of spontaneous circulation (ROSC).^22,23,25–29^ (c) Five studies reported survival to hospital discharge.^22,23,27–29^ (d) Four studies reported survival with good neurological outcome.^22,23,27,29^ (e) Only two studies reported post-ROSC arterial blood gas parameters, specifically pO₂ and pCO₂ levels.^24,26^

This variability in outcome availability led to different total sample sizes across the respective meta-analyses.

### 3.2 Study Characteristics

Table 1 shows the study characteristics of the included studies. Extracted variables included first author, publication year, study design, country, setting (in-hospital vs out-of-hospital cardiac arrest), method of CPR (manual vs mechanical), ventilation mode or strategy, sample size, and outcomes measured. Extracted outcomes included *return of spontaneous circulation (ROSC)*, *survival to hospital admission or discharge*, *neurological recovery*, and *physiologic parameters* such as arterial or venous blood gases, ETCO₂ , and SpO₂ . Any discrepancies were resolved by consensus with the third reviewer (A.R.).

**Table 1.** Characteristics of Included Studies. Table 1 summarizes the baseline characteristics of the included studies, including study design, sample size, intervention arms, ventilation strategies, and outcomes measured.

| Author (Year) | Design | Country | Sample Size (Total / MV / Manual) | Ventilator Strategy | Setting | CPR Method | Outcomes Measured |
| --- | --- | --- | --- | --- | --- | --- | --- |
| Kim et al, 2025 | Retrospective case-control with propensity matching | Republic of Korea | 522 (261 / 261) | VCV | OHC A | Mechanical | ROSC, Survival to admission, Survival to discharge, Neurological outcome |
| Lofti et | RCT | Iran | 61 (31 | VCV | IHCA | Manual | VBG |
| **al, 2025** |  |  | / 30) |  |  |  | parameter<br>s (PaO<sub>2</sub> ,<br>pH,<br>PaCO<sub>2</sub> ,<br>HCO<sub>3</sub><sup>-</sup>),<br>ETCO<sub>2</sub> ,<br>SpO<sub>2</sub> |
| **Malinver**<br>**ni et al,**<br>**2024** | Retrospec<br>tive multicentr<br>e | Belgiu<br>m | 2,566<br>(298 /<br>2,268) | Non-<br>synchroni<br>zed,<br>BiPAP<br>mode | OHC<br>A | Both | ROSC,<br>Favorable<br>neurologic<br>al<br>outcome |
| **Shin et**<br>**al, 2024** | RCT | Korea | 60 (30<br>/ 30) | VCV | OHC<br>A | Manual | ROSC,<br>sustained<br>ROSC ≥<br>20 min,<br>ABG<br>parameter<br>s,<br>ventilation<br>and lung<br>injury<br>scores |
| **Tangpais**<br>**arn et al,**<br>**2023** | RCT | Thaila<br>nd | 60 (30<br>/ 30) | VCV | IHCA | Manual | ABG<br>parameter<br>s, ROSC,<br>Survival to<br>discharge,<br>Neurologic<br>al<br>outcome,<br>Complicati<br>ons |
| **Turowski**<br>**et al,**<br>**2025** | Retrospec<br>tive cross-<br>sectional | Germa<br>ny | 3,192<br>(2,140<br>/<br>1,055) | IPPV,<br>CCSV,<br>BiPAP | OHC<br>A | Both | ROSC,<br>Survival to<br>discharge |
| **SYMEVE**<br>**CA 1,**<br>**2023** | Prospectiv<br>e quasi-<br>experimen | Spain | 150<br>(71 /<br>79) | IPPV | OHC<br>A | Manual | ROSC,<br>Time to<br>advanced |
|  | tal |  |  |  |  |  | CPR,<br>Survival,<br>Neurologic<br>al<br>outcome |
| **SYMEVE**<br>**CA 2,**<br>**2025** | Prospectiv<br>e quasi-<br>experimen<br>tal | Spain | 521<br>(245 /<br>276) | IPPV,<br>CCSV | OHC<br>A | Manual | ROSC,<br>Hospital<br>survival,<br>Neurologic<br>al<br>outcome |

RCT – Randomized Controlled Trail, VCV – Volume Controlled Ventilation, BiPAP – Bilevel Positive Airway Pressure, IPPV – Intermittent Positive Pressure Ventilation, CCSV – Chest Compression Synchronized Ventilation, OHCA – Out of Hospital Cardiac Arrest, IHCA – In-Hospital Cardiac Arrest, ROSC – Return of Spontaneous Circulation, VBG – Venous Blood Gas, ABG – Arterial Blood Gas, CPR – Cardiopulmonary Resuscitation.

The eight included studies encompassed a total of 5,130 participants, comprising both randomized controlled trials and observational studies conducted between 2023 and 2025 across multiple countries (Korea, Iran, Belgium, Thailand, Germany, and Spain). Of these, eight studies (n = 5,130) were included in the qualitative synthesis, and seven five and four studies were eligible for quantitative meta-analysis of primary outcomes namely ROSC, survival to hospital discharge and survival with good neurological outcomes.

Study designs included three randomized controlled trials, two prospective quasi-experimental studies, and three retrospective cohort studies (including one multicenter registry). The interventions predominantly used *volume-controlled ventilation (VCV)* or *intermittent positive pressure ventilation (IPPV)*, while comparators employed standard *bag-valve-mask (BVM)* manual ventilation.

Mechanical ventilation protocols typically used a tidal volume of 6–7 mL/kg ideal body weight (approximately 500 mL for men, 400–450 mL for women), a respiratory rate of 10–12 breaths per minute, and a fraction of inspired oxygen (FiO2) of 1.0 (100%), with positive end-expiratory pressure (PEEP) most commonly set at 0–5 cmH2O. Pressure settings reported included peak inspiratory pressures up to 45–60 cmH2O and inspiratory time of 1 second with an I:E ratio of 1:1–1:5. CPR was delivered manually in most studies, though two incorporated mechanical chest compression devices.

### 3.3 Risk of Bias

Supplemental Figure S1 summarizes the results of the risk-of-bias assessments for the included studies. Among the three randomized controlled trials (RCTs), all were judged to have low risk of bias across all five domains (randomization process, deviations from intended interventions, missing outcome data, outcome measurement, and selective reporting). Accordingly, the overall risk of bias for these RCTs was rated as *low*.

In contrast, the five non-randomized studies demonstrated a high risk of bias in the confounding domain (Domain 1), primarily due to unmeasured or uncontrolled variables such as time to initiation of CPR, CPR quality, use of advanced airway devices, and availability of mechanical compression or defibrillation resources. Four of these studies also exhibited moderate or serious concerns in *participant selection (Domain 2)* and *intervention deviation (Domain 4)*, reflecting potential baseline imbalances and differences in ventilation protocols. Bias related to *classification of interventions (Domain 3)*, *missing data (Domain 5)*, and *outcome measurement (Domain 6)* was generally low. *Selective reporting bias (Domain 7)* was low across all studies. Overall, the non-randomized evidence was of moderate-to-serious risk of bias, primarily driven by confounding and selection domains.

### 3.4 Primary Outcomes

#### 3.4.1 Return of Spontaneous Circulation (ROSC)

##### Randomized Controlled Trials (RCTs)

Two RCTs (n = 120 participants) reported ROSC outcomes (Figure 2a). Pooled analysis using a random-effects REML model showed no statistically significant difference between mechanical ventilation and manual ventilation during CPR (OR 1.49; 95% CI 0.73–3.07; p = 0.27; I² = 0%). Although mechanical ventilation showed a numerical trend toward improved ROSC, the limited sample size and low event rates likely contributed to the lack of statistical significance.

**Figure 2a.**
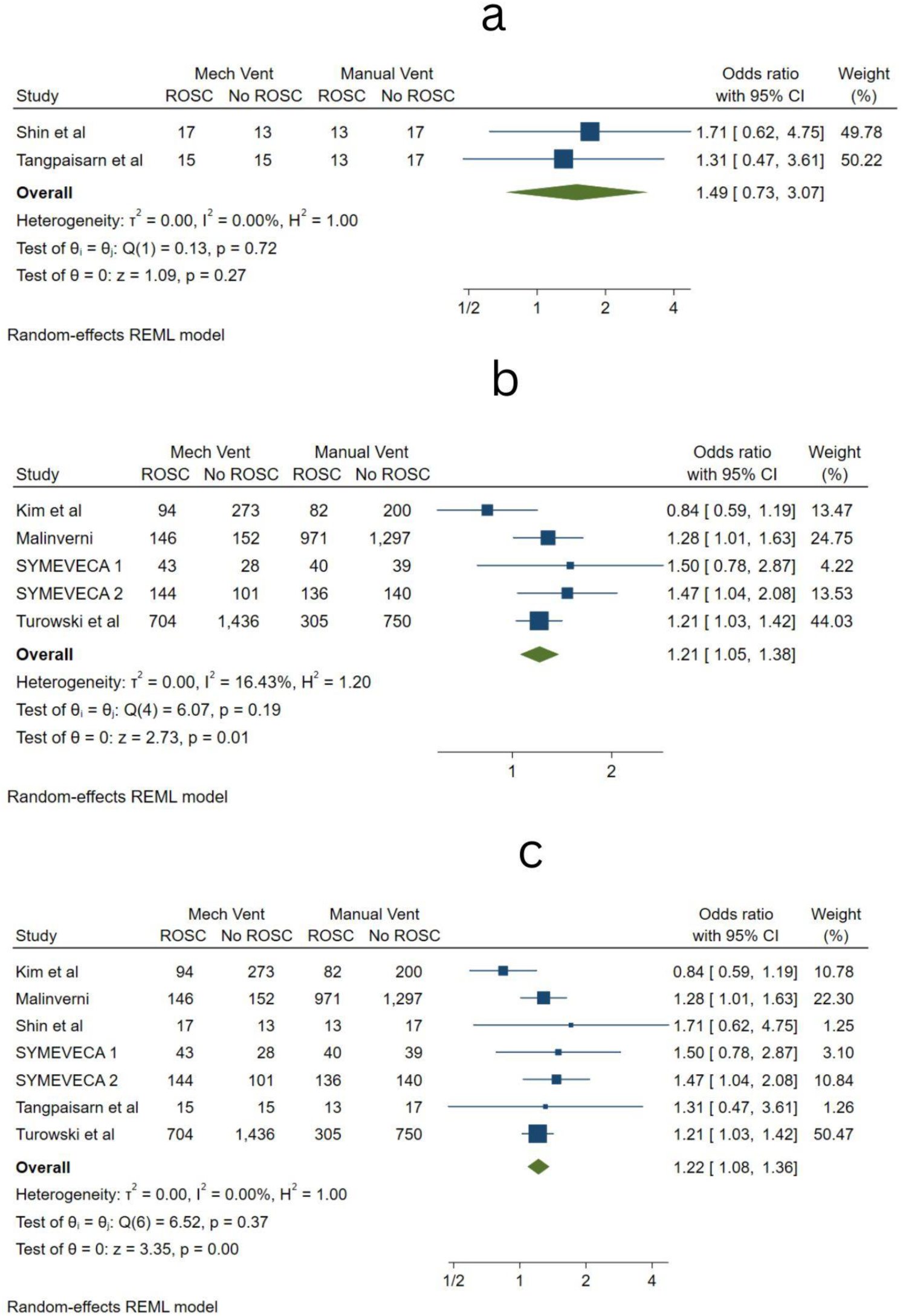
Forest plot of two randomized controlled trials (n=120) comparing mechanical ventilation with manual ventilation during CPR. Pooled odds ratio (OR 1.49; 95% CI 0.73–3.07) shows a non-significant trend favoring mechanical ventilation, with low heterogeneity (I²=0%). **Figure 2b**. Forest plot of five observational studies (n=7,081). Mechanical ventilation was associated with higher ROSC (OR 1.21; 95% CI 1.05–1.38; p=0.01), with low heterogeneity (I²=16.4%). **Figure 2c**. Pooled forest plot including seven studies (n=7,201) comparing mechanical vs manual ventilation. Mechanical ventilation improved ROSC (OR 1.22; 95% CI 1.08–1.36; p<0.001) with no observed heterogeneity (I²=0%).

### Non-Randomized Studies (nRCTs)

Five observational and quasi-experimental studies (n = 7,081 participants) evaluated ROSC. Mechanical ventilation was associated with a statistically significant increase in ROSC compared with manual ventilation (OR 1.21; 95% CI 1.05–1.38; p = 0.01; I² = 16.43%) (Figure 2b). The mild heterogeneity likely reflects variations in arrest setting, EMS systems, and ventilation strategies.

### Overall (Combined) Analysis

Seven studies (n = 7,201 participants) reported ROSC outcomes. Pooled analysis using a random-effects model demonstrated that mechanical ventilation during CPR improved ROSC compared with manual ventilation (OR 1.22; 95% CI 1.08–1.36; p = 0.00; I² = 0%) (Figure 2c). The low heterogeneity indicates consistent findings across diverse study designs and settings.

### Subgroup Analysis by Study Setting (IHCA vs OHCA)

A prespecified subgroup analysis assessed whether the effect of ventilation strategy differed based on the cardiac arrest setting.

- In-Hospital Cardiac Arrest (IHCA)

Only one study reported IHCA-exclusive results, demonstrating no significant difference between mechanical and manual ventilation (OR 1.31; 95% CI 0.47–3.61).

- Out-of-Hospital Cardiac Arrest (OHCA)

Six studies reported OHCA data, showing a significant improvement in ROSC with mechanical ventilation (OR 1.22; 95% CI 1.05–1.41; P = 0.009; I² = 23%).

The test for subgroup differences was not statistically significant (P = 0.89), suggesting that any apparent difference between settings is likely attributable to sample size variation rather than a true interaction.

### Bayesian Meta-Analysis

Bayesian model averaging showed anecdotal evidence in favour of a non-zero effect (posterior probability 62.3%; inclusion BF = 1.65). The pooled posterior mean log OR was 0.12 (95% CrI, 0.00–0.32), corresponding to an OR of 1.13 (95% CrI, 1.00–1.37). This suggests a small possible improvement in ROSC, although the credible interval includes no effect. There was no evidence for between-study heterogeneity (posterior probability 46.4%; inclusion BF = 0.87). The 95% prediction interval (OR 0.70–1.59) indicates considerable uncertainty and suggests that future studies may plausibly observe benefit, no effect, or harm (Figure 3).

**Figure 3.**
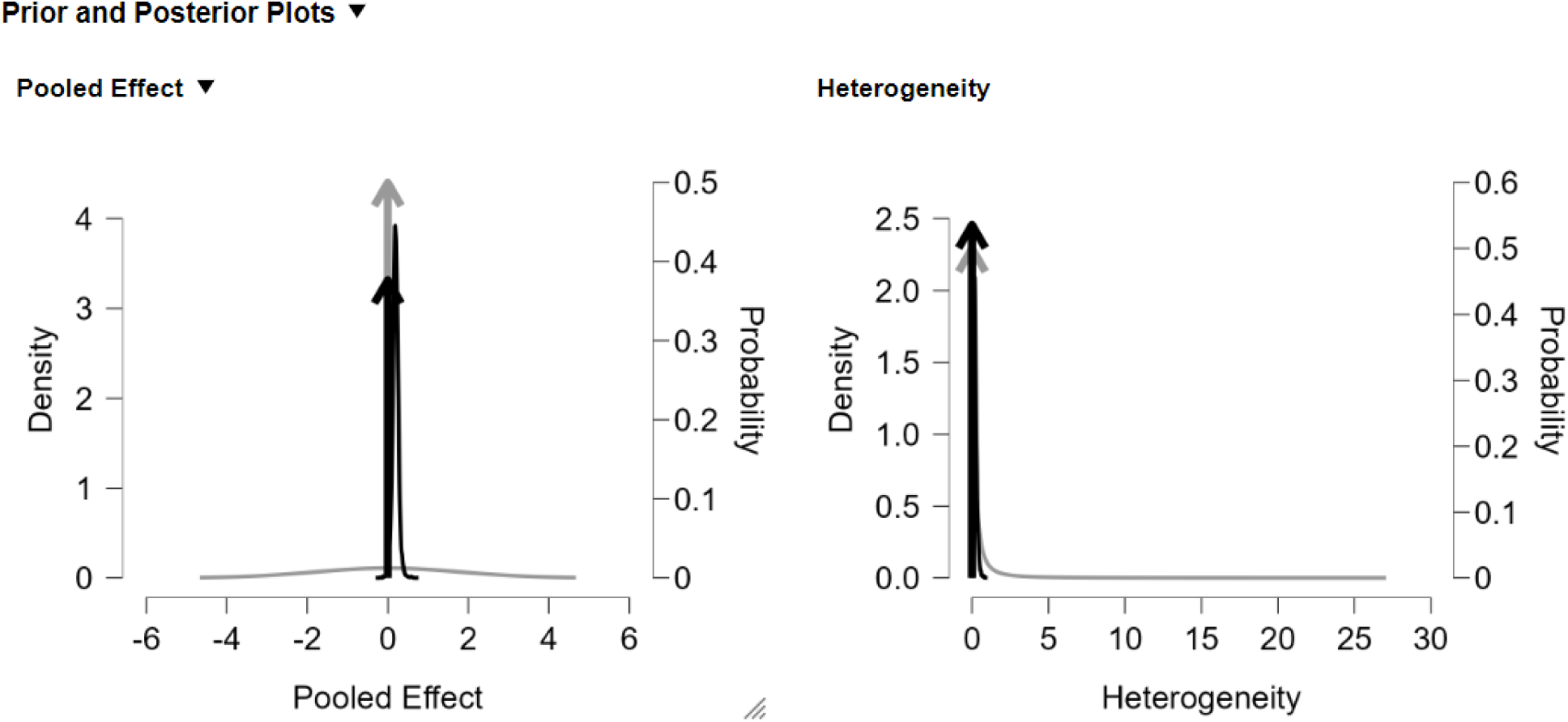
Graphical comparison of prior and posterior distributions for the pooled log odds ratio and between-study heterogeneity. Posterior mean log OR was 0.12 (95% CrI 0.00–0.32), corresponding to OR 1.13 (95% CrI 1.00–1.37).

#### 3.4.2 Survival to Hospital Discharge

One randomized controlled study and four non-randomized studies (n = 4,575 participants) reported survival to discharge (Figure 4). Mechanical ventilation during CPR resulted in higher survival to discharge compared with manual ventilation (OR 1.34; 95% CI 1.05–1.71; p = 0.02; I² = 0%). The absence of heterogeneity (τ² = 0.00) indicates highly consistent findings across diverse populations and EMS systems.

**Figure 4.**
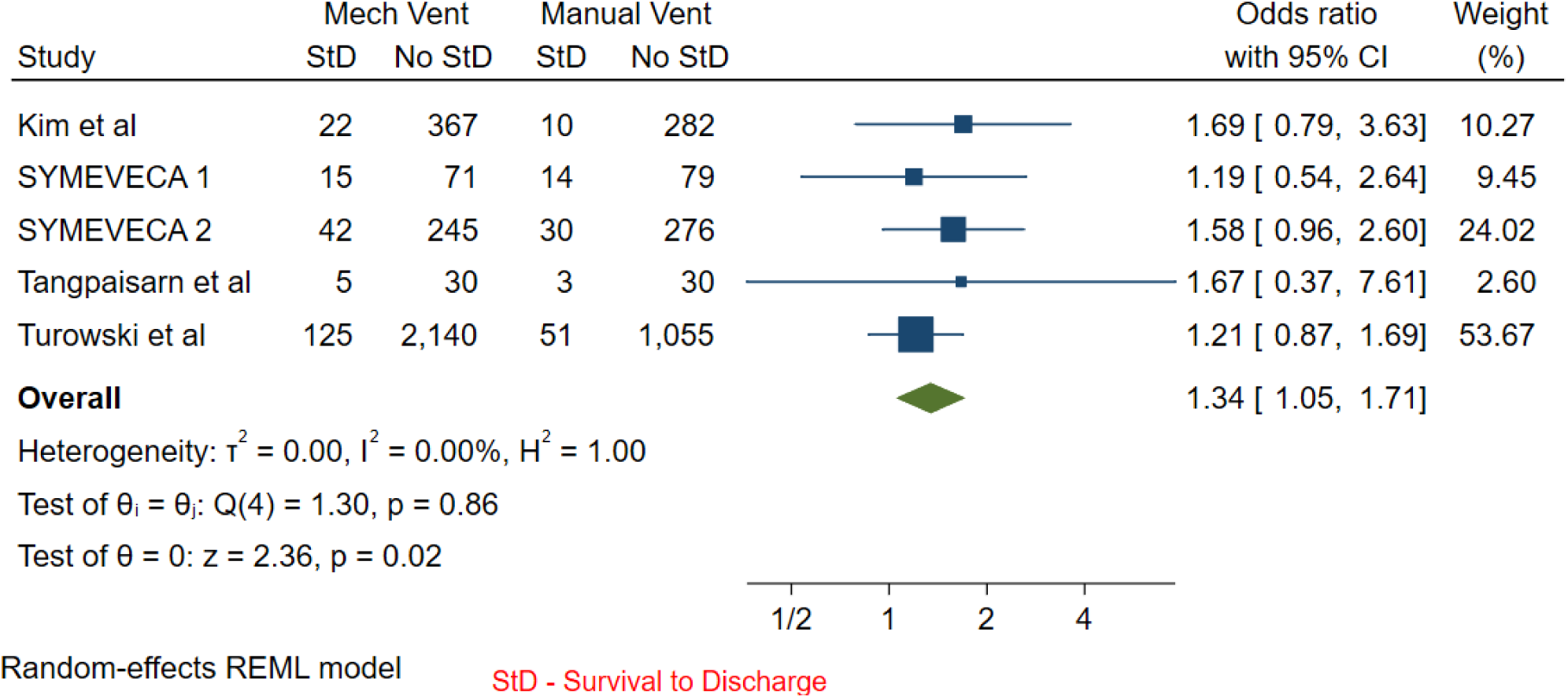
Forest plot of one randomized controlled trial and four observational studies (n=4,575). Mechanical ventilation improved survival to discharge (OR 1.39; 95% CI 1.08–1.77; p=0.02), with no heterogeneity (I²=0%).

#### 3.4.3 Survival with Good Neurological Outcome

One randomized controlled study and three studies (n = 1,380 participants) assessed favorable neurological recovery (CPC 1–2) (Figure 5). Mechanical ventilation improved neurological outcomes (OR 1.54; 95% CI 1.00–2.36; P = 0.05; I² = 0%). These findings support the physiological plausibility that controlled ventilation may enhance post-ROSC cerebral perfusion.

**Figure 5.**
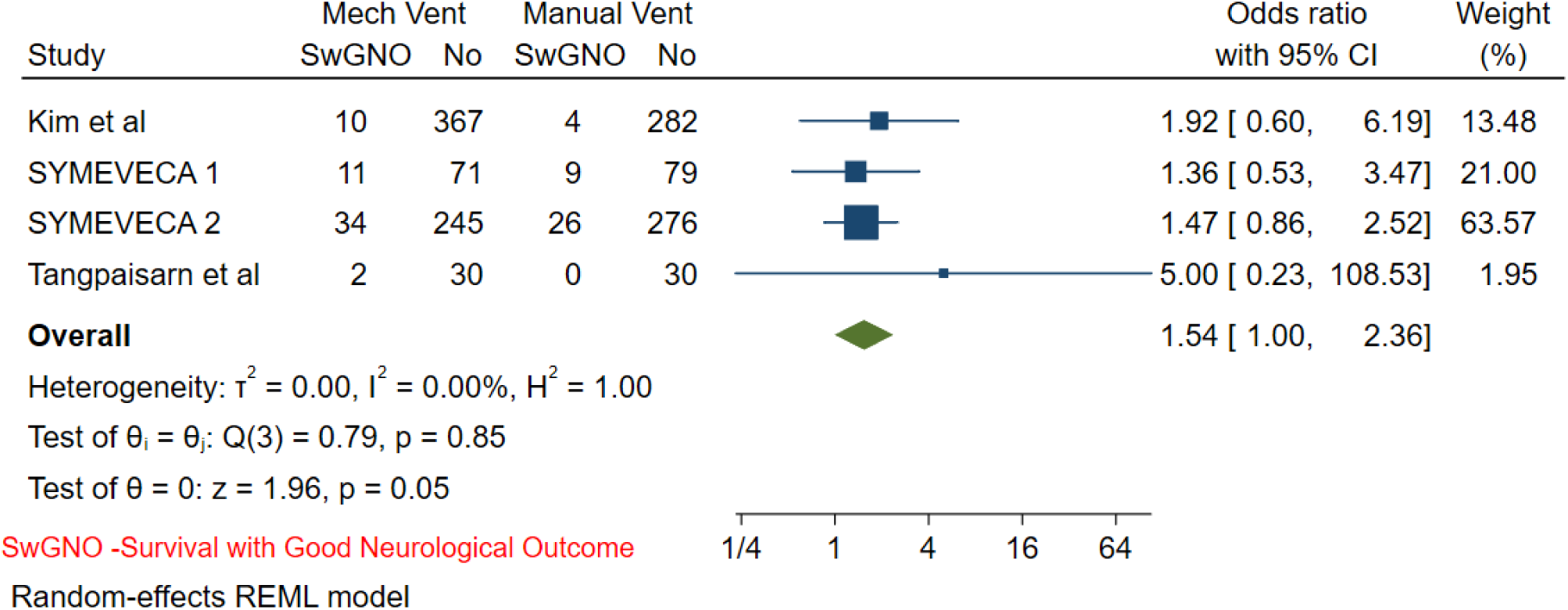
Forest plot of four studies (n=1,380). Mechanical ventilation improved neurological outcomes (OR 1.54; 95% CI 1.00–2.36; p=0.05), with no heterogeneity (I²=0%).

**Figure 6.**
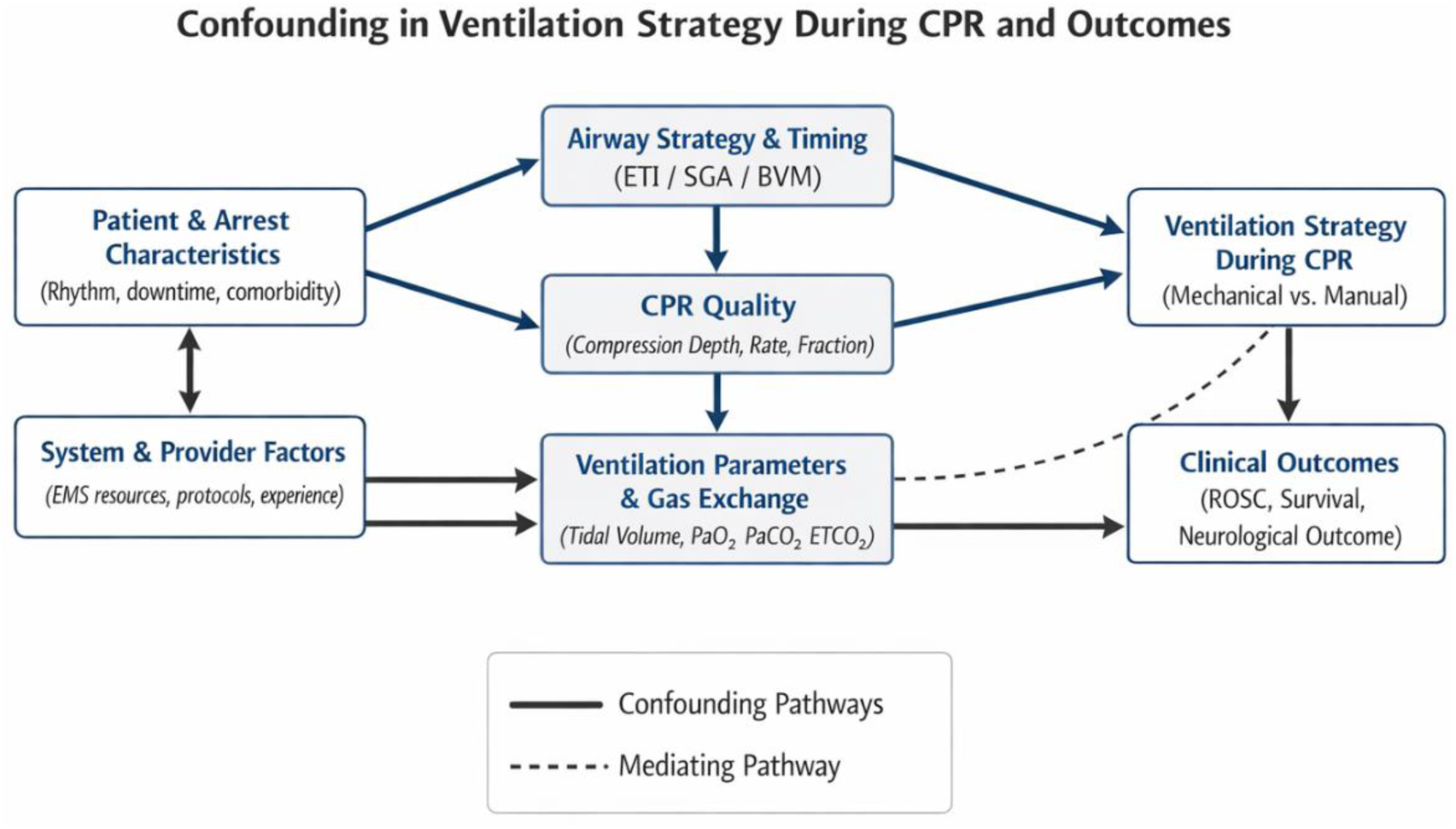
Directed Acyclic Graph. This DAG depicts the hypothesized causal structure linking ventilation strategy during CPR to clinical outcomes. Mechanical ventilation is conditionally dependent on advanced airway placement, which is influenced by patient and arrest characteristics, clinician decision-making, and system-level factors. Airway strategy therefore represents a major confounder, as it is causally related to both the exposure (mechanical vs manual ventilation) and outcomes (return of spontaneous circulation, survival to hospital discharge, and favorable neurological outcome). Additional confounding arises from CPR quality and resuscitation system factors, which may influence both ventilation strategy and outcomes. Improved gas exchange during CPR is shown as a potential mediating pathway between ventilation strategy and outcomes; however, this pathway cannot be isolated in predominantly observational data. The DAG illustrates why observed associations should be interpreted as non-causal and hypothesis-generating.

### 3.5 Secondary Outcomes

#### Post-ROSC Arterial Blood Gas Parameters

##### pO₂ Levels

Two RCTs (n = 121 participants) reported post-ROSC pO₂ . Mechanical ventilation resulted in higher pO₂ than manual ventilation (MD 15.31 mmHg; 95% CI 10.31–19.96; P < 0.0001; I² = 0%) (Supplemental Figure S3).

##### pCO₂ Levels

The same two RCTs also reported pCO₂ values. Mechanical ventilation produced higher pCO₂ compared with manual ventilation (MD 12.98 mmHg lower; 95% CI 6.77–19.47; P < 0.00001; I² = 19.47%) (Supplemental Figure S4). These findings explain that manual ventilation causes hyperventilation and a resulting carbon di oxide washout. However, the high carbon di-oxide also suggests that the recommended respiratory rate of 10 breaths per minute needs to be introspected as it causes hypercapnia when applied very strictly.

### 3.6 GRADE Certainty of Evidence

According to the GRADE assessment, the overall certainty of evidence was low for clinical outcomes and moderate for physiological outcomes (Table 2). The certainty for ROSC, survival to discharge, and neurological recovery was downgraded primarily due to the predominance of non-randomized studies, introducing a serious risk of bias from confounding and selection effects. Although the pooled estimates demonstrated consistent findings with low statistical heterogeneity, the evidence was further downgraded for imprecision in effect estimates—particularly within the smaller RCTs—and for possible publication bias, suggested by asymmetry in the funnel plots. In contrast, outcomes related to post-ROSC arterial blood gas parameters (pO₂ and pCO₂), derived exclusively from RCTs, were rated as moderate certainty, with only one level of downgrading for imprecision. Overall, the GRADE evaluation indicates that while mechanical ventilation during CPR is associated with improved outcomes, the confidence in these findings is limited and underscores the need for larger, high-quality randomized trials.

**Table 2.**
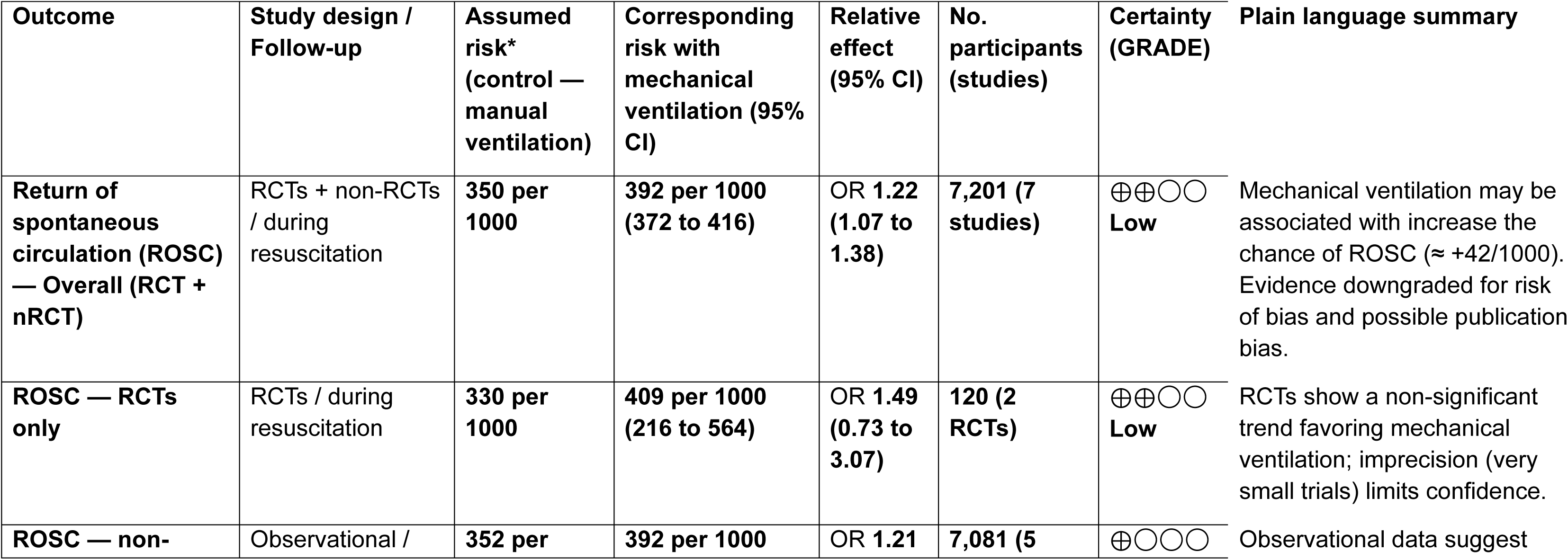
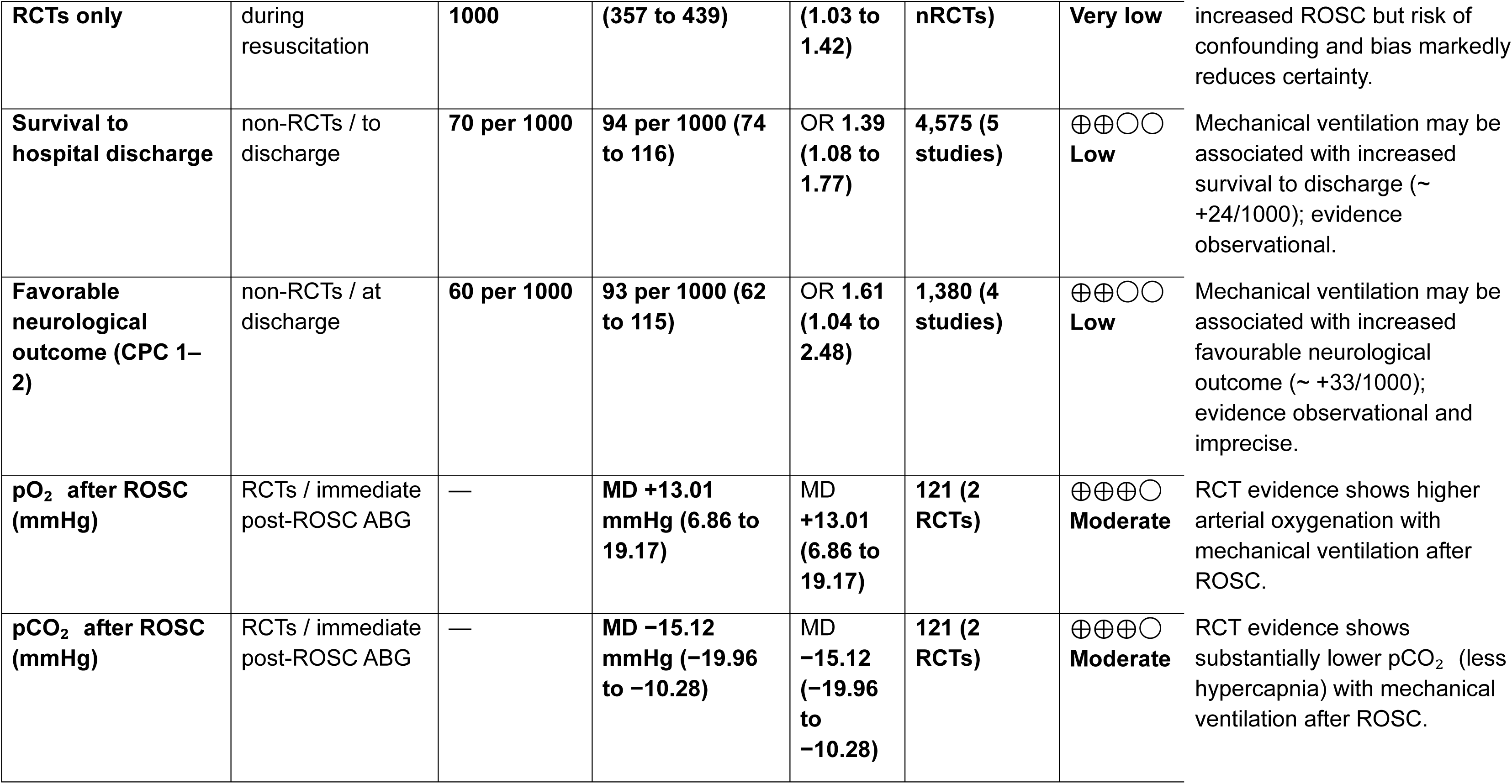
GRADE Summary of Findings for Mechanical vs Manual Ventilation During CPR.

## 4 DISCUSSION

This systematic review and meta-analysis provide the most comprehensive synthesis to date comparing mechanical and manual ventilation during adult cardiopulmonary resuscitation. Across eight studies involving more than five thousand patients, mechanical ventilation was associated with higher rates of return of spontaneous circulation, improved survival to hospital discharge, and better neurological outcomes, along with more consistent post-resuscitation arterial blood gas profiles. However, these associations must be interpreted with caution because the evidence base is characterized by three major sources of uncertainty: airway-related structural confounding, dominance of observational evidence, and limited, underpowered randomized trial data.

First, mechanical ventilation during CPR is structurally dependent on advanced airway placement. In contemporary practice, mechanical ventilation is almost universally applied after endotracheal intubation or supraglottic airway insertion, whereas manual ventilation is frequently delivered via bag-valve-mask. As a result, ventilation modality is not independently assigned but conditional on airway strategy. Airway management itself influences outcomes through multiple pathways, including effects on intrathoracic pressure, ventilation–compression coordination, and resuscitation workflow. This creates structural confounding, with airway strategy acting as a common cause of both the exposure and outcomes, thereby violating the exchangeability assumption required for unbiased causal inference in observational studies.

Second, most included evidence is observational, and patient selection further amplifies confounding. The decision to intubate during CPR is not random and is often influenced by anticipated resuscitation duration, perceived physiologic reserve, and clinician judgment regarding the likelihood of survival. Patients receiving mechanical ventilation may therefore represent a subgroup selected for more aggressive or prolonged resuscitation, whereas patients remaining on bag-valve-mask ventilation may have had more severe physiologic compromise or earlier termination of efforts. In this context, mechanical ventilation may function as a marker of resuscitation intensity rather than a direct determinant of improved outcomes, biasing effect estimates toward apparent benefit. Additionally, key confounders such as intubation timing, CPR quality metrics, and airway-related interruptions were inconsistently reported and rarely adjusted for.

Third, randomized evidence remains limited and underpowered. Although the available trials demonstrate more consistent ventilation and gas exchange with mechanical ventilation, they are small, focus largely on physiologic outcomes, and lack sufficient power to detect differences in patient-centered endpoints. As a result, randomized data are insufficient to confirm or refute the associations observed in larger observational cohorts. The predominance of observational evidence therefore constrains the certainty of pooled estimates despite low statistical heterogeneity.

Within these limitations, the observed physiologic differences between ventilation strategies remain clinically relevant but should be interpreted cautiously. Manual bag-valve ventilation is prone to variability in tidal volume and respiratory rate, while mechanical ventilation may provide greater consistency. However, because these physiologic effects cannot be isolated from airway strategy and resuscitation context in observational data, they should be viewed as potential mediators rather than established causal mechanisms (Figure 16).

Operational feasibility further limits generalizability. Ventilator–compression interactions, pressure-limit alarms, and triggering failures can compromise breath delivery during CPR, particularly in volume-controlled modes. Prehospital environments introduce additional constraints related to device portability, power supply, transport logistics, and provider training. These considerations underscore that ventilation modality is one component within a broader resuscitation system and that physiologic advantages alone are insufficient to justify widespread adoption.

In summary, mechanical ventilation during CPR is associated with improved clinical and physiologic outcomes, but these findings are embedded within a complex causal structure dominated by airway strategy, observational confounding, and limited randomized evidence. Ventilation modality should therefore be considered an adjunct within a multifactorial resuscitation process rather than an independent therapeutic intervention. The current evidence should be regarded as hypothesis-generating and highlights the need for adequately powered randomized trials that explicitly address airway timing, ventilation strategy, and CPR quality.

### Strengths and Limitations

This review synthesizes the available randomized and observational evidence comparing mechanical and manual ventilation during adult CPR using a comprehensive search strategy, prespecified protocol registration, and stratified analyses by study design. Clinically relevant outcomes, including neurologic status, were evaluated, and evidence certainty was transparently assessed using the GRADE framework. Bayesian analyses complemented conventional random-effects models by explicitly characterizing uncertainty. Several limitations warrant consideration. Randomized evidence remains sparse and underpowered, and most outcome estimates are derived from observational studies susceptible to confounding, particularly related to airway strategy and resuscitation intensity.

Heterogeneity in ventilation modes, airway practices, arrest settings, and EMS systems complicates direct comparisons. Analyses were limited to study-level data, precluding adjustment for key prognostic variables such as intubation timing and CPR quality. The small number of studies also limited formal assessment of publication bias.

### Future Directions

Future research should prioritize adequately powered randomized trials that disentangle ventilation modality from airway strategy and resuscitation intensity. Such studies should incorporate standardized reporting of airway timing, CPR quality metrics, ventilation parameters, and system-level characteristics. Pragmatic investigations are also needed to address operational feasibility, including ventilator–compression interactions, device optimization, workflow integration, and provider training, particularly in prehospital environments.

## 5. Conclusion

Mechanical ventilation during CPR is associated with higher rates of return of spontaneous circulation, survival, and favorable neurological outcomes, as well as more consistent oxygenation and ventilation parameters. However, these associations arise predominantly from observational studies in which mechanical ventilation is structurally linked to advanced airway placement and other determinants of resuscitation intensity, introducing substantial potential for confounding. The randomized evidence remains limited, and the overall certainty of evidence is low. Consequently, no causal conclusions can be drawn. Mechanical ventilation may represent a promising approach to standardizing intra-arrest ventilation, but its clinical role remains uncertain and should be considered hypothesis-generating pending confirmation in rigorously designed randomized trials.

## Data Availability

Data included in this review is already avaialble as only published materials were used

## Presented in Meetings

No

## Prior Publication

No

## Source of Funding

None

## Disclosure

None

## Conflicts of interest

None

## Data Sharing

Data will be shared by the corresponding author upon reasonable request.

## Author’s Contribution

**GR,** – Conceptualization, Methodology, Software, Validation, Formal analysis, Investigation, Data Curation, Writing – Original Draft, Visualization, Supervision, Project administration.

**SM, AR –** Methodology, Software, Formal analysis, Data curation.

**GP, EG, VV, PR, HR -** Writing - Review and Editing.

---

**Supplemental Figure S1.**
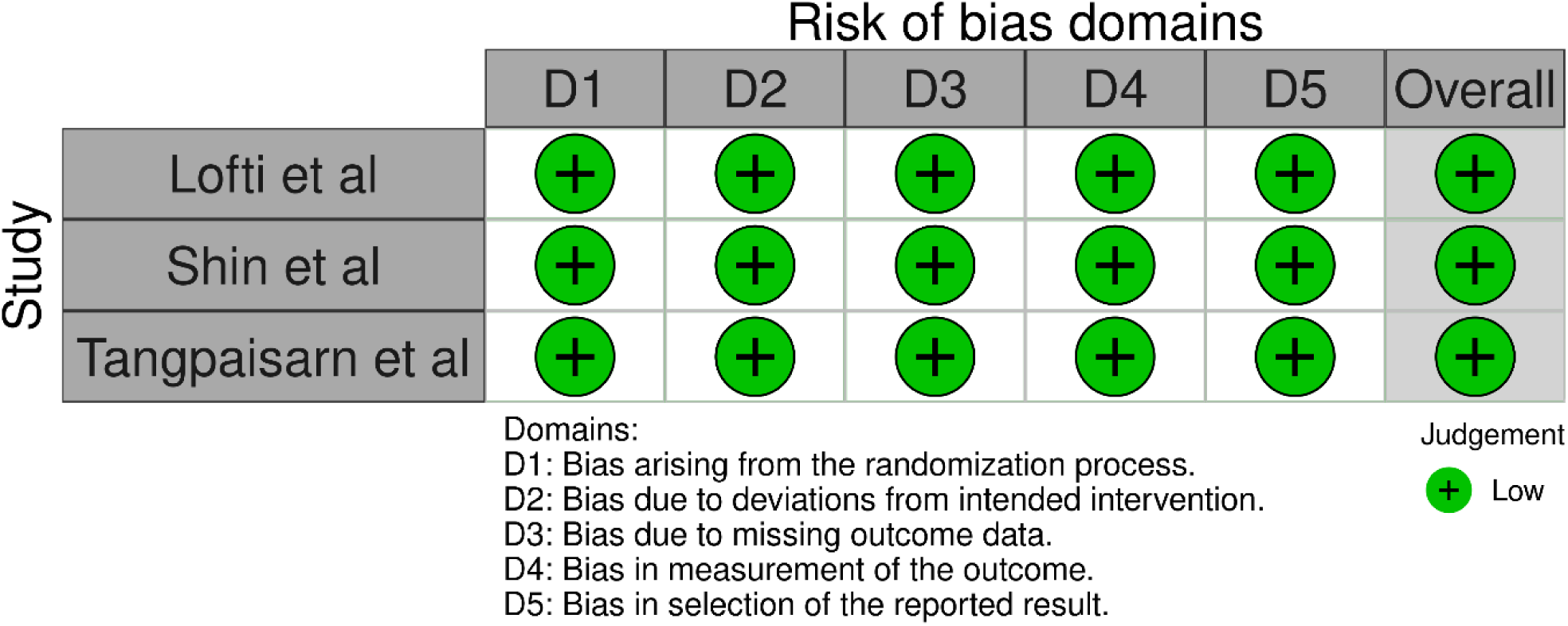
Risk of Bias Assessment for Randomized Controlled Trials (RoB 2 Tool) Traffic-light and summary plots illustrating domain-level and overall risk-of-bias judgments for each RCT.

**Supplemental Figure S2.**
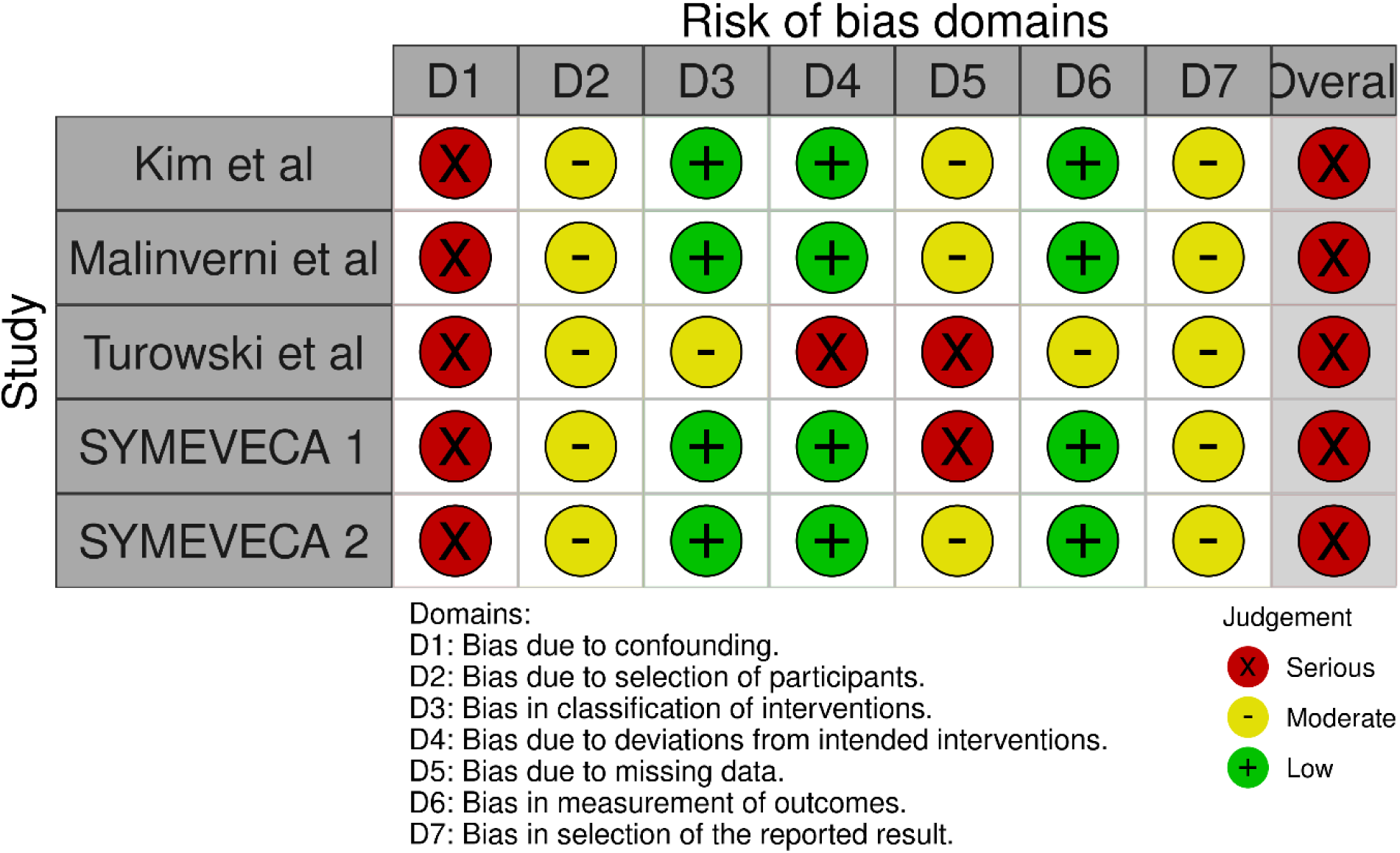
Risk of Bias Assessment for Observational Studies (ROBINS-I Tool) Domain-specific and overall risk-of-bias profiles for the five non-randomized studies, highlighting serious confounding risk.

**Supplemental Figure S3.**
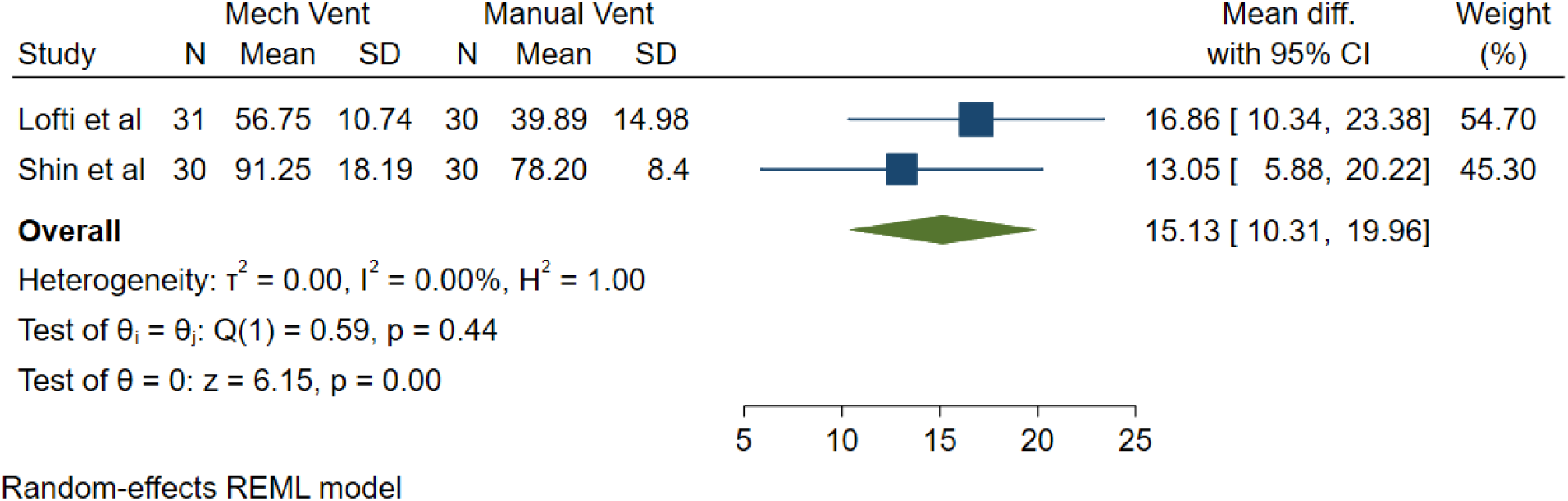
Post-ROSC Arterial Oxygen Tension (pO₂) — Randomized Trials. Forest plot of two RCTs (n=121) showing higher pO₂ in patients ventilated mechanically (mean difference +15.31 mmHg; 95% CI 10.31–19.96; p<0.0001; I²=0%).

**Supplemental Figure S4.**
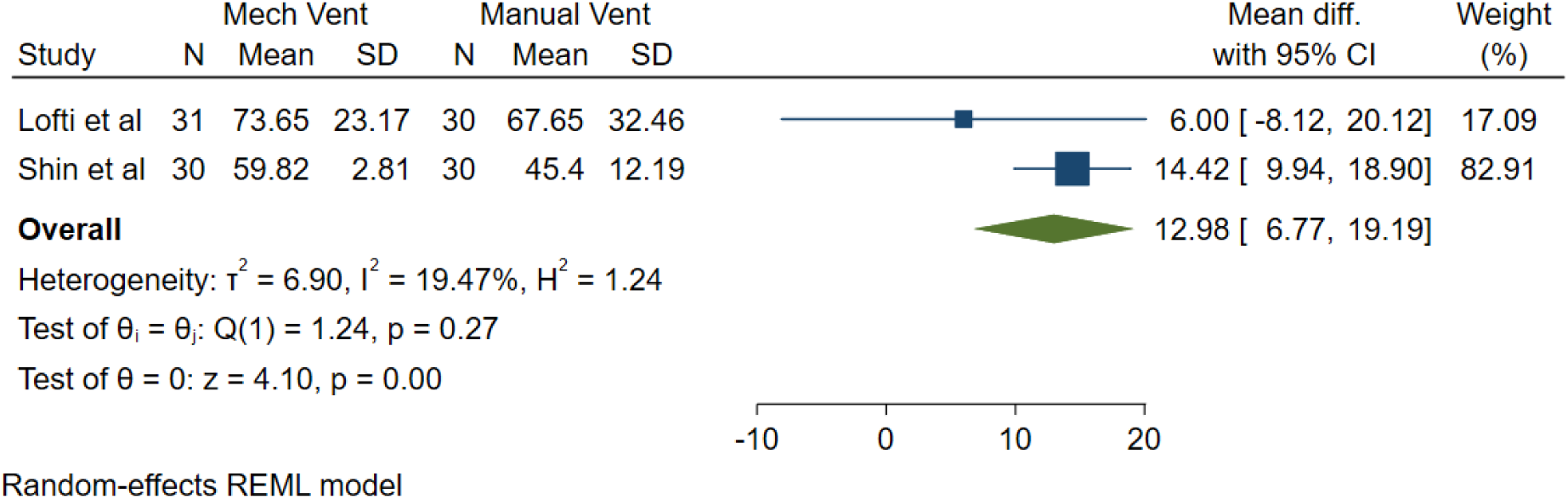
Post-ROSC Arterial Carbon Dioxide Tension (pCO₂) — Randomized Trials. Forest plot showing lower pCO₂ (reduced hyperventilation washout) with mechanical ventilation compared with manual bag ventilation (mean difference −12.98 mmHg; 95% CI −19.47 to −6.77; p<0.00001; I²=19%).

**Supplemental Table S1.** Full Electronic Search Strategy.

|  |
| --- |
| PubMed<br>"respiration, artificial"[MeSH Terms] AND ("Heart Arrest"[MeSH Terms] OR "Out-of-Hospital Cardiac Arrest"[MeSH Terms]) |
| EMBASE<br>'mechanical ventilation during cpr' OR (mechanical AND ('ventilation'/exp OR ventilation) AND during AND cpr) |
| SCOPUS<br>TITLE-ABS-KEY ( Mechanical ventilation during CPR ) |

## Notes

### Competing Interest Statement

The authors have declared no competing interest.

### Author Declarations

This is a systematic review and meta-analysis and hence IRB approval is not required

